# Elevated plasma levels of CXCL16 in severe COVID-19 patients

**DOI:** 10.1101/2021.09.07.21263222

**Authors:** Sandra P. Smieszek, Vasilios M. Polymeropoulos, Christos M. Polymeropoulos, Bartlomiej P. Przychodzen, Gunther Birznieks, Mihael H. Polymeropoulos

## Abstract

Genome-wide association studies have recently identified 3p21.31, with lead variant pointing to the *CXCR6* gene, as the strongest thus far reported susceptibility risk locus for severe manifestation of COVID-19. In order the determine its role, we measured plasma levels of Chemokine (C□X□C motif) ligand 16 (CXCL16) in the plasma of COVID-19 hospitalized patients. CXCL16 interacts with CXCR6 promoting chemotaxis or cell adhesion. The CXCR6/CXCL16 axis mediates homing of T cells to the lungs in disease and hyper-expression is associated with localised cellular injury. To characterize the CXCR6/CXCL16 axis in the pathogenesis of severe COVID-19, plasma concentrations of CXCL16 collected at baseline from 115 hospitalized COVID-19 patients participating in ODYSSEY COVID-19 clinical trial were assessed together with a set of controls. We report elevated levels of CXCL16 in a cohort of COVID-19 hospitalized patients. Specifically, we report significant elevation of CXCL16 plasma levels in association with severity of COVID-19 (as defined by WHO scale) (*P*-value<0.02). Our current study is the largest thus far study reporting CXCL16 levels in COVID-19 hospitalized patients (with whole-genome sequencing data available). The results further support the significant role of the CXCR6/CXCL16 axis in the immunopathogenesis of severe COVID-19 and warrants further studies to understand which patients would benefit most from targeted treatments.

## Introduction

Genome-wide association studies have recently identified 3p21.31, encompassing the *CXCR6* gene, as the strongest thus far reported susceptibility region associated with a severe manifestation of COVID-19 (rs73064425, OR=2.14, *P*-value=4.77E-30)^1^. Another lead severity variant rs10490770:T>C pointing to *CXCR6* was also reported with a *P*-value of 1.443E-73 (hospitalization) and 2.20E-61 (critical illness) by the COVID-19 Host Genetics Initiative^2^. Nakanishi et al., reported that risk allele (rs10490770) carriers experienced an increased risk of COVID-19 related mortality (HR 1.5, 95% CI 1.3-1.8)^3^. Risk allele carriers had increased odds of several COVID-19 complications: severe respiratory failure (OR 2.0, 95% CI 1.6-2.6), venous thromboembolism (OR 1.7, 95% CI 1.2-2.4)^3^.

Chemokine (C□X□C motif) ligand 16 (CXCL16) is synthesized as a transmembrane molecule that is expressed as a cell surface-bound molecule, and as a soluble chemokine^4^. CXCL16 interacts with CXCR6 in leukocytes and other cells promoting chemotaxis or cell adhesion. It has been shown that inflammatory cytokines such as IFNγ and TNFα promote CXCL16 expression^5^. CXCL16 has been previously implicated in the pathogenesis of lung injury upon which it is released and functions as a chemoattractant for CXCR6+ T, natural killer (NK), B, and dendritic cells^6^. CXCL16 levels were elevated in the serum of acute lung injury patients in comparison to controls^6^. CXCR6 is expressed on T cells not only in T helper type 1 (Th1) inflammation but also in Th2 inflammation, where it is increased after allergen challenge. It was previously shown that reduction in CD8+ T-lymphocytes occurred between C57BL/6 and CXCR6KO mice^7^. CXCR6 is a major coreceptor of HIV type 2 variants^8^. The CXCR6/CXCL16 axis mediates homing of T cells to the lungs in disease and when hyper expressed is associated with localized cellular injury^7^. This CXCR6/CXCL16 axis is involved in lung pathology also associated with other infections, including influenza. Antagonism was reported to result in reduced inflammation^6^. In a recent report authors have shown thatknockdown ofCXCR6 reversed the effect of CXCL16 and so did the treatment with the p38 inhibitor SB203580 abolished the effects of CXCL16^6^.Another recemt study has shown that the absence of CXCR6 significantly decreases airway CD8 resident memory T cells due to altered trafficking of CXCR6−/− cells within the lung^9^, Moreover mice lacking CXCL16 also had decreased CD8 resident memory T cells in the airways and ultimately blocking CXCL16 resulted in the inhibition of the steady-state maintenance of airway resident memory T cells^9^.

## Methods

### Participants

ODYSSEY is a double-blinded Phase 3 study with a planned randomization of a total of 300 hospitalized severely ill COVID-19 patients (Clinicaltrials.gov: NCT04326426). Inclusion criteria for the study comprised of: 1. Adults aged 18-90; 2. confirmed laboratory COVID-19 infection; 3. confirmed pneumonia by chest radiograph or computed tomography; 4. fever defined as temperature ≥ 36.6 °C armpit, ≥ 37.2 C oral, or ≥ 37.8 °C rectal since admission or the use of antipyretics; 5. PaO2 / FiO2 ≤ 300; 6. In patient hospitalization. Patients were to be followed for up to 28 days to record clinical outcomes. Patients’ clinical progress was recorded on a 7 point clinical status ordinal scale defined as follows: 1-Death; 2-Hospitalized on mechanical ventilation or ECMO; 3-Hospitalized on non invasive ventilation or high-flow oxygen supplementation; 4-Hospitalized requiring supplemental oxygen; 5-Hospitalized not requiring supplemental oxygen, requiring continued medical care; 6-Hospitalized not requiring supplemental oxygen, not requiring continued medical care; 7-Not hospitalized. Main Exclusion Criteria included: 1. Inability to provide informed consent or to have an authorized relative or designated person provide informed consent, or to comply with the protocol requirements; 2. Known allergy to tradipitant or other neurokinin-1 antagonists; 3. Pregnancy; 4. Uncontrolled HIV, HBV, or HCV infection; 5. Other uncontrolled medically significant diseases; 6. Enrollment in another clinical trial of an investigational therapy; 7. Alanine aminotransferase > 5X Upper Limit of Normal or Creatinine clearance < 50 m 174 l / min; 8. Requiring mechanical ventilation for > 72 hours.

### CXCL16 measurement

To characterize the CXCR6/CXCL16 axis in the pathogenesis of severe COVID-19, plasma concentrations of CXCL16 from 115 hospitalized COVID-19 patients participating in ODYSSEY COVID-19 clinical trial ((NCT04326426)) and additionally samples from 37 controls were assessed. CXCL16 levels in plasma were determined with an enzyme□linked immunosorbent assay (ELISA) assay (<u>Thermofisher</u> ID: # EHCXCL16) performed on plasma collected at baseline in cases and in a set of healthy controls (assay range 2.74-2000 pg/mL).

### Genetic Analysis

Incoming nucleic acid samples are quantified using fluorescent-based assays (PicoGreen) to accurately determine whether sufficient material is available for library preparation and sequencing. DNA sample size distributions are profiled by a Fragment Analyzer (Advanced Analytics) or BioAnalyzer (Agilent Technologies), to assess sample quality and integrity. HumanCoreExome 24v1.3 array was performed on all human DNA samples sequenced. Whole genome sequencing (WGS) libraries were prepared using the Truseq DNA PCR-free Library Preparation Kit. Whole Genome data were processed on NYGC automated pipeline. Paired-end 150 bp reads were aligned to the GRCh37 human reference (BWA-MEM v0.7.8) and processed with GATK best-practices workflow (GATK v3.4.0). The mean coverage was 35.8, it reflects the samples average. All high quality variants obtained from GATK were annotated for functional effects (intronic, intergenic, splicing, nonsynonymous, stopgain and frameshifts) based on RefSeq transcripts using Annovar31. Additionally, Annovar was used to match general population frequencies from public databases (Exac, gnomAD, ESP6500, 1000g) and to prioritize rare, loss-of-function variants. Linear models adjusted for PC, age and sex were conducted in PLINK.

## Results

We report elevated levels of CXCL16 in a set of COVID-19 hospitalized patients. Baseline demographics of the studied cohort are presented in **Table 1**. Clinical charactestics are provided in **Table 2**. Specifically, we report significantly elevated levels of CXCL16 in a subset of COVID-19 severe (as defined by WHO scale) hospitalized patients ((*P*-value<0.02) displayed on **Figure 1**). The most significant difference reported in plasma CXCL16 between most severe (WHO-2) COVID-19 hospitalized cases and less severe patients (*P*-value<0.02) as well as most severe and controls. We furthermore report a larger variance within the cases, signifying further necessity of stratification based on clinical characteristics (ANCOVA p-value – 0.001, covariates: age, sex, BMI). The effect is not driven by age nor BMI, or sex (Figure 2 – Supplemental Material). We further investigated the correlation between IFN-γ, TNF-α. We report lack thereof at baseline (Figure 3 – Supplemental Material). We were able to replicate the lead variant association (rs10490770) in our cohort of hospitalized patients (cases n 140 control n 1900) with confirmed COVID-19 infection (OR 1.9, CI 1.2-3.5, *P*-value <0.002). The carriers of this variant were at higher risk, of being in the hospitized severe group (as defined by the WHO scale), finding which is consistent across multiple COVID-19 GWAS results reported thus far. Moreover, we inspected the association between the lead variant status and directly CXCL16 plasma levels. This association between lead coding variant carrier status and CXCL16 levels did not attain statistical significance (result that is trending), a finding that warrants further replication in a larger sample size. FYCO1 variant within the extended haplotype did attain statistical significance (*P*-value <0.0001) and the result is displayed in Supplementary Figure 4.

**Table 1.**

| **Characteristic** | **Total**<br>**(N=141)** |
| --- | --- |
| **Age (years)** | **64 (54 - 72)** |
| **Sex, n (%)** |  |
| Male | 91 (64.5) |
| Female | 50 (35.5) |
| **Race, n (%)** |  |
| Asian | 2 (1.4) |
| Black or African American | 35 (24.8) |
| Native Hawaiian or Other Pacific Islander | 1 (0.7) |
| White | 73 (51.8) |
| Other | 30 (21.3) |
| **Any Comorbidities, n (%)** | **135 (95.7)** |
| Hypertension | 80 (56.7) |
| Diabetes | 56 (39.7) |
| Coronary Heart Disease | 10 (7.1) |
| Asthma | 16 (11.3) |
| **7 Point Ordinal Scale at Baseline, n (%)** |  |
| 2 - Hospitalized on mechanical ventilation or ECMO | 8 (5.7) |
| 3 - Hospitalized on non-invasive ventilation or high-flow oxygen supplement | 69 (48.9) |
| 4 - Hospitalized requiring supplemental oxygen | 61 (43.3) |
| 5 - Hospitalized not requiring supplemental oxygen, requiring continued medical care | 3 (2.1) |

**Table 2.**
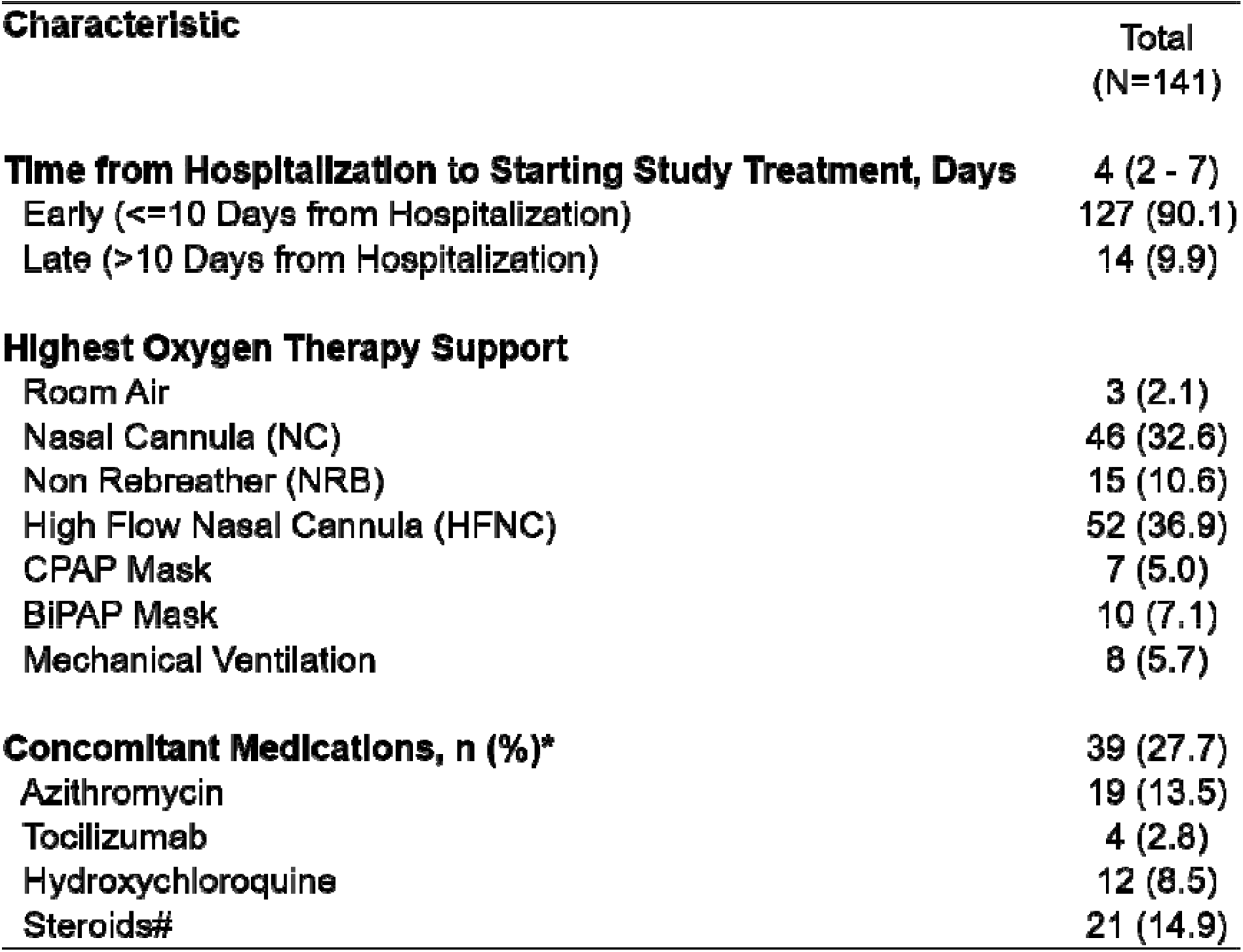

**Figure 1.**
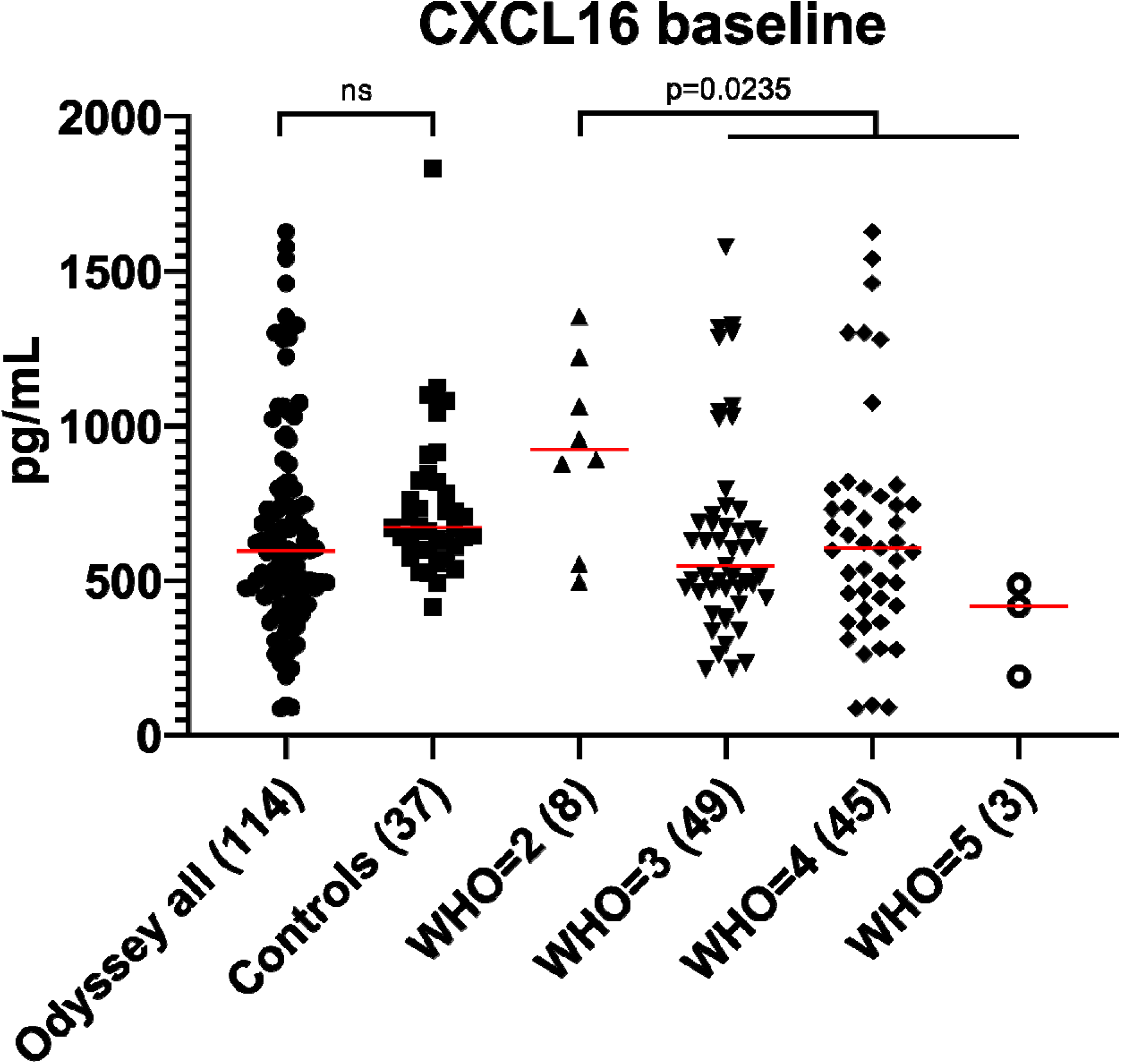
Baseline plasma CXCL16 levels in association with severity of COVID-19 manifestation and case control analysis. Severe (as defined by WHO scale) cases had significantly higher levels of CXCL16 – effect that is statistically significant.

## Discussion

Previously it was demonstrated that circulating CD8+CXCR6+ T cells were significantly elevated with advanced age, yet virtually absent in patients with severe COVID-19. In a smaller sample set, plasma levels of CXCL16 were significantly upregulated in severe COVID-19 patients compared to mild COVID-19 patients^10^. Differential expression of *CXCR6* and *CXCL16* mRNA was observed in severe COVID-19 compared to mild disease and significant functional polymorphisms in *CXCR6* are linked to viral control^1^.

Furthermore, based on lung tissue autopsy data, the top differentially expressed genes from GWAS hits included *FYCO1*, specifically in AT2 cells, ciliated cells, and club cells^11^. FYCO1 coding variant (rs33910087) is an eQTL for CXCR6, whereas CXCR6 was differentially expressed in lung CD8 T cells in COVID-19 infected lung tissue^11^.

Prior studies studies targeting the axis (both knockout and pharmacological blocking) support the hypothesis that antagonism may constitute potential therapies^6^,^9^. It may be the case that the treatment of the hyper-expression of the CXCL16/CXCR6 axis hence reduction of the severely heighteed homing signal may elicit such therapeutic effects, for example in a cytokine storm. Besides the potential implications in lung injury, both the CXCR6 and its ligand plays an important role in mediating a proinflammatory microenvironment for tumor growth in hepatocellular carcinoma^12^. Here targeting the CXCR6/CXCL16 axis is needed to elucidate the mechanism whereby neutrophils are affected in the tumor environment^12^. Authors propose one such orally bioavailable compound - antagonist of the CXCR6 receptor shown to decrease growth in a mouse xenograft model of hepatocellular carcinoma^12^. Further studies on antagonism of the axis are needed to explain the timing of such antagonism treatment that would result in optimal outcomes. Our current study, is the largest thus far reporting on the plasma levels of CXCL16 in association with severity in COVID-19 patients. The results support the significant role of the CXCR6/CXCL16 axis in the immunopathogenesis of severe COVID-19.Further studies warrant analysis of outcomes based on the carrier status for this locus and on the levels of CXCL16.

## Data Availability

Data available upon request

---

**Supplemental Figure 2.**
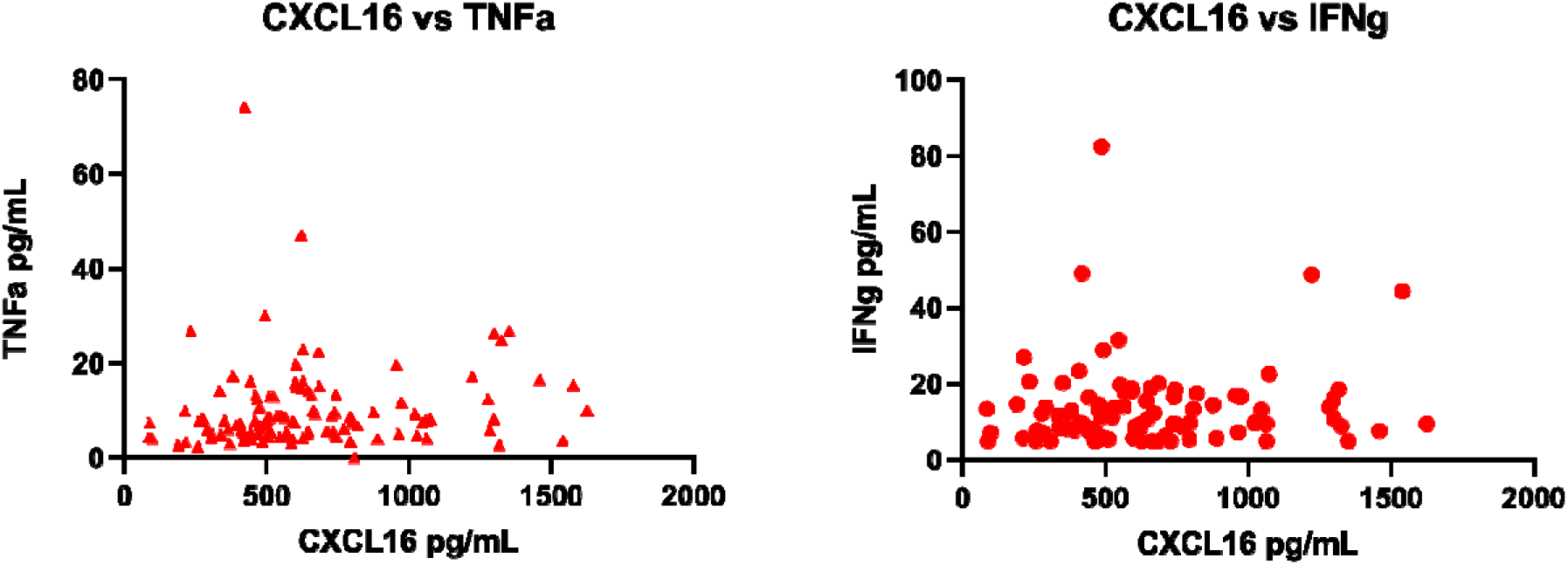
Scatter plot between CXCL16 and IFNg and TNFa (baseline samples collected from COVID-19 hospitlized patients with severity determined with WHO scale)

**Supplemental Figure 3.**
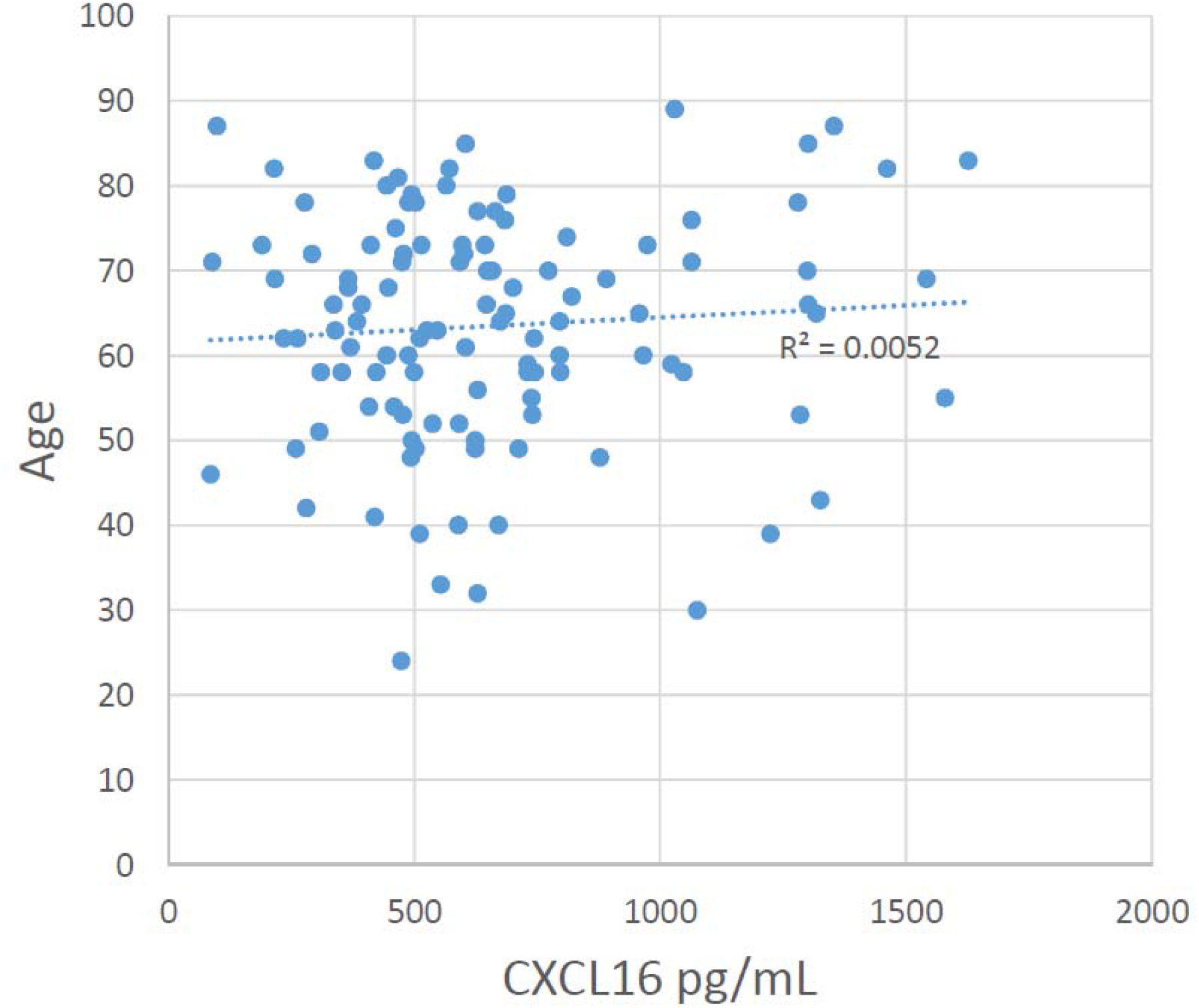
Correlation between age and CXCL16 plasma levels (collected at baseline form COVID-19 hospilized patients)

**Supplemental Figure 4.**
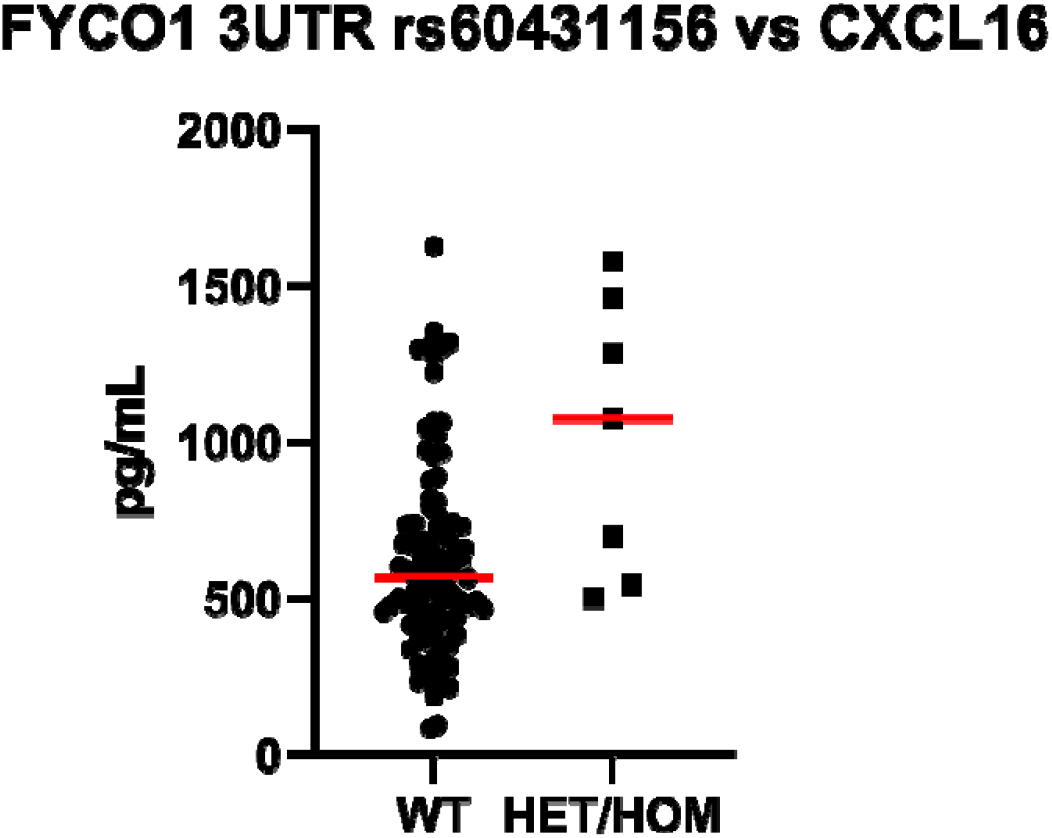
Association between genotype status and CXCL16 plasma levels (collected at baseline form COVID-19 hospilized patients)

